# Seventeen-Year National Pain Prevalence Trends Among U.S. Military Veterans

**DOI:** 10.1101/2023.03.27.23287408

**Authors:** Kenneth Adam Taylor, Flavia Penteado Kapos, Jason Arthur Sharpe, Andrzej Stanislaw Kosinski, Daniel I. Rhon, Adam Payne Goode

**Affiliations:** Department of Orthopaedic Surgery, Duke University School of Medicine, Durham, North Carolina; Duke Clinical Research Institute, Duke University School of Medicine, Durham, North Carolina; Center for Child Health, Behavior and Development, Seattle Children’s Research Institute, Seattle, Washington; Center of Innovation to Accelerate Discovery and Practice Transformation (ADAPT), Durham Veterans Affairs (VA) Healthcare System, Durham, North Carolina; Department of Biostatistics and Bioinformatics, Duke University School of Medicine, Durham, North Carolina; Department of Rehabilitation Medicine, School of Medicine, Uniformed Services University of the Health Sciences, Bethesda, Maryland; Department of Population Health Sciences, Duke University School of Medicine, Durham, North Carolina

## Abstract

**Importance:** U.S. military veterans experience higher pain prevalence and severity than nonveterans. However, it is unclear how these differences have changed over time. Previous studies are limited to veterans receiving care from the Veterans Health Administration.

**Objective:** To characterize pain prevalence trends in the overall population of U.S. veterans compared to nonveterans, using nationally-representative data.

**Design:** Repeated cross-sectional study. Data: National Health Interview Survey, 2002-2018. Analysis: January 2023.

**Setting:** Population-based survey of noninstitutionalized U.S. adults.

**Participants:** Across the 17-year period, mean annual weighted population was 229.7 million adults (unweighted sample total: n=506,639; unweighted sample annual mean: n=29,802).

**Exposure:** Veteran status.

**Main Outcomes:** Crude and demographics-adjusted pain prevalence trend differences between veterans and nonveterans across five pain variables (severe headache or migraine, facial pain, neck pain, low back pain, and joint pain) and two composite variables (any pain [≥1 prevalent pain] and multiple pains [≥2 prevalent pains]).

**Results:** Weighted proportion of veterans varied from 11.48% in 2002 (highest) to 8.41% in 2017 (lowest). Across the study period, crude prevalence was generally similar or higher among veterans than nonveterans for all pain variables except for severe headache or migraine and facial pain. When equalizing age, sex, race, and ethnicity, pain prevalence among veterans remained similar or higher than nonveterans for all pain variables. From 2002 to 2018 there was an absolute increase (95% CI) in pain prevalence among veterans (severe headache or migraine: 2.0% [1.6% to 2.4%]; facial pain: 1.9% [1.4% to 2.4%]; neck pain: 4.7% [4.1% to 5.2%]; joint pain: 11.4% [10.8% to 11.9%]; low back pain: 10.3% [9.5% to 11.1%]; any pain: 10.0% [9.6% to 10.4%]; and multiple pains: 9.9% [9.2% to 10.6%]. Crude and adjusted analyses indicated prevalence of all pain variables increased more among veterans than nonveterans from 2002 to 2018.

**Conclusion and Relevance:** Veterans had similar or higher adjusted prevalence and higher rates of increase over time for all pain variables compared to nonveterans. Continued pain prevalence increase among veterans may impact healthcare utilization (within and outside of the VHA), underscoring the need for improved pain prevention and care programs for these individuals with disproportionate pain burden.

**Key Points:** *Question:* How have differences in pain prevalence between U.S. military veterans and nonveterans changed from 2002 to 2018?

*Findings:* In this repeated cross-sectional study using 17 years of nationally-representative data of U.S. adults, we found similar or higher pain prevalence and higher prevalence increase over time among veterans than nonveterans, even when accounting for differing distributions of age, sex, race, and ethnicity.

*Meaning:* Disparities in pain prevalence between veterans and nonveterans worsened from 2002 to 2018, underscoring the need for improved prevention and pain care programs for veterans, who experience a disproportionate burden of pain.

## INTRODUCTION

The U.S. population included 19.4 million U.S. military veterans in 2020^1^ – individuals who served in any military service branch but are no longer serving – approximately 6.9% to 7.7% of the U.S. population since 2011.^2,3^ Veterans are disproportionately impacted by multiple chronic health conditions and severe psychological distress compared to age-matched nonveteran U.S. adults^4,5^ and more likely to report worse overall health and health-related quality of life.^6-8^ They also have higher prevalence of painful conditions than nonveterans, such as doctor-diagnosed arthritis,^6,8,9^ and self-reported severe pain.^10^

Pain prevalence is trending upward among U.S. adults overall. A study using Medical Expenditure Panel Survey (MEPS) data from 1997 to 2014 reported care-seeking or disability episodes related to painful conditions increased from 32.9% to 41.1% among U.S. adults.^11^ Another study using National Health Interview Survey (NHIS) data from 2002 to 2018 reported increasing prevalence of each available pain location across a wide range of sociodemographic subgroups among U.S. adults aged 25-84.^12^

Pain prevalence is also trending upward in veteran-specific populations. Veterans Health Administration (VHA) data from 2000 to 2007 shows a relative 39.7% increase in low back pain (LBP) care.^13^ Additionally, 2004 to 2011 VHA neck and spinal pain prevalence increased from 1.9% to 2.5% and 2.6% to 4.2%, respectively.^14^ While these trends are informative, they are limited to VHA-enrolled veterans, who are generally less healthy than veterans receiving care outside the VHA.^15,16^ Therefore, these findings are not generalizable to the broader U.S. veteran population and provide limited ability to compare trends to the overall U.S. adult population.

Therefore, our goal was to characterize pain prevalence trends in the overall population of U.S. veterans compared to nonveterans, using nationally representative data.

## METHODS

### Study Design and Target Population

This study used data from the NHIS Sample Adult and Person questionnaires from 2002 to 2018.^17^ Additional details regarding sampling design are available from the National Center for Health Statistics (NCHS).^18^ We present additional NHIS design and target population information relevant to our study in the **eMethods**. The NCHS Research Ethics Review Board reviews and approves NHIS content and methods annually. Interviewers obtain verbal consent for participation from all survey respondents.

### Veteran Status

Our primary exposure of interest was veteran status (veterans vs. nonveterans). Since 2011, the NHIS identifies veterans by asking, “Did you ever serve on active duty in the U.S. Armed Forces, military Reserves, or National Guard?” Before 2011, the NHIS identified veterans by asking, “Have you ever been honorably discharged from active duty in the U.S. Army, Navy, Air Force, Marine Corps, or Coast Guard?;” as a result, National Guard or Reserves veterans nor veterans discharged for misconduct (e.g., general or other than honorable discharge) or court-martial (e.g., bad conduct or dishonorable discharge) were not included.

### Pain Prevalence

The NHIS asks sample adults about pain (yes/no) in four regions that lasted a day or more over the past 3 months (excluding “aches and pains that are fleeting or minor”): severe headache or migraine, facial pain, neck pain, and LBP. Respondents who report experiencing LBP are asked a follow-up question regarding the presence of associated leg pain that “spread down either leg to areas below the knee.” An additional question is asked about joint pain (excluding pain in the joints of the back or neck) in the past 30 days. The structure and wording of these questions (**eTable 1**) is consistent across all NHIS years used in this study.

Using the five pain variables above, we created two non-mutually exclusive variables. First, we created an “any pain” variable, identifying individuals with ≥1 prevalent pain variable. Second, we created a “multiple pains” variable, identifying individuals with ≥2 prevalent pain variables. While the NHIS provided data regarding pain interference (2016-2017), pain intensity, chronicity of pain, and pain management (2015-2018), we did not include these variables in our analysis because they were not available for our entire study period.^19^

### Statistical Methods

We calculated the crude and adjusted national prevalence (95% confidence intervals [CIs]) of each pain variable among veterans and nonveterans for each quarter from 2002 through 2018^20-22^ using SAS 9.4 (Cary, NC). RStudio was used to visualize trends over time by veteran status.^23-25^ Analyses incorporated primary sampling units, stratum, and sample adult weights to account for the NHIS design complexity. Sample adult weights account for sample selection probability adjusted for household non-response, age, race, and sex using quarterly Census Bureau population control totals.^17^

Crude estimates represent observable national-level trends in veterans and non-veterans. Adjusted prevalence estimates allow more direct comparison between these two groups by standardizing on demographic characteristics. We used respondent-reported age, sex, race, and Hispanic ethnicity as model covariates to account for demographic characteristic differences between veterans and nonveterans. We describe the NHIS coding of demographic characteristic covariates in the **eMethods**.

We used linear regression with a time-by-veteran-status multiplicative interaction term to calculate mean difference in prevalence change over time in each group and associated 95% CIs using robust standard errors. We used annualized sample adult weighted prevalence estimates for 2002 and 2018 to calculate the total absolute change (prevalence_2018_– prevalence_2002_) and relative change ([prevalence_2018_–prevalence_2002_]/prevalence_2002_) in pain prevalence across the entire 17-year study period for both point estimates and 95% CIs. Based on the extreme rarity of missingness for both veteran status and pain variables, we did not apply any imputation approaches; we excluded individuals missing exposure or outcome data. There were no missing data for age and sex, and missing data in race and Hispanic ethnicity are imputed by NCHS before public data release.^26^

## RESULTS

The mean weighted population between 2002 and 2018 was 229.7 million noninstitutionalized U.S. adults. This ranged from 205.8 million (in 2002) to 249.5 million (in 2018). Veterans accounted for between 8.4% (95% CI: 8.0%, 8.8%; 2017) and 11.5% (95% CI: 11.1%, 11.9%; 2002) of the weighted adult population across the study period (**eTable 2**). Veteran status was missing for a small proportion (≤0.35% [95% CI: 0.21%, 0.48%] annually) of the weighted population due to refusal to answer, failure to ascertain, or respondent (or their proxy) being unsure of veteran status (**eTable 3**). We present a summary of weighted covariates by year for veterans in **eTable 4** and nonveterans in **eTable 5**. Missingness of pain variables was rare (**eTable 6, eTable 7, eTable 8, eTable 9, eTable 10, eTable 11, eTable 12**). We present the absolute and relative change in annual prevalence of each pain variable at the start (2002) and end (2018) of the study period in **eTable 13** (veterans) and **eTable 14** (nonveterans).

### Severe Headache or Migraine

Crude prevalence of severe headache or migraine (**Figure 1**) was consistently lower among veterans, ranging from 6.1% (95% CI: 4.1%, 8.1%; 2006, 2^nd^ quarter) to 12.4% (95% CI: 10.1%, 14.6%; 2011, 2^nd^ quarter) among veterans and from 12.3% (95% CI: 11.3%, 13.3%; 2007, 2^nd^quarter) to 17.7% (95% CI: 16.7%, 18.7%; 2010, 1^st^ quarter) among nonveterans. Prevalence estimates and trend lines closely approximated when adjusting for demographic differences (**eFigure 1**), with veterans having slightly higher prevalence of severe headache or migraine on average.

**Figure 1.**
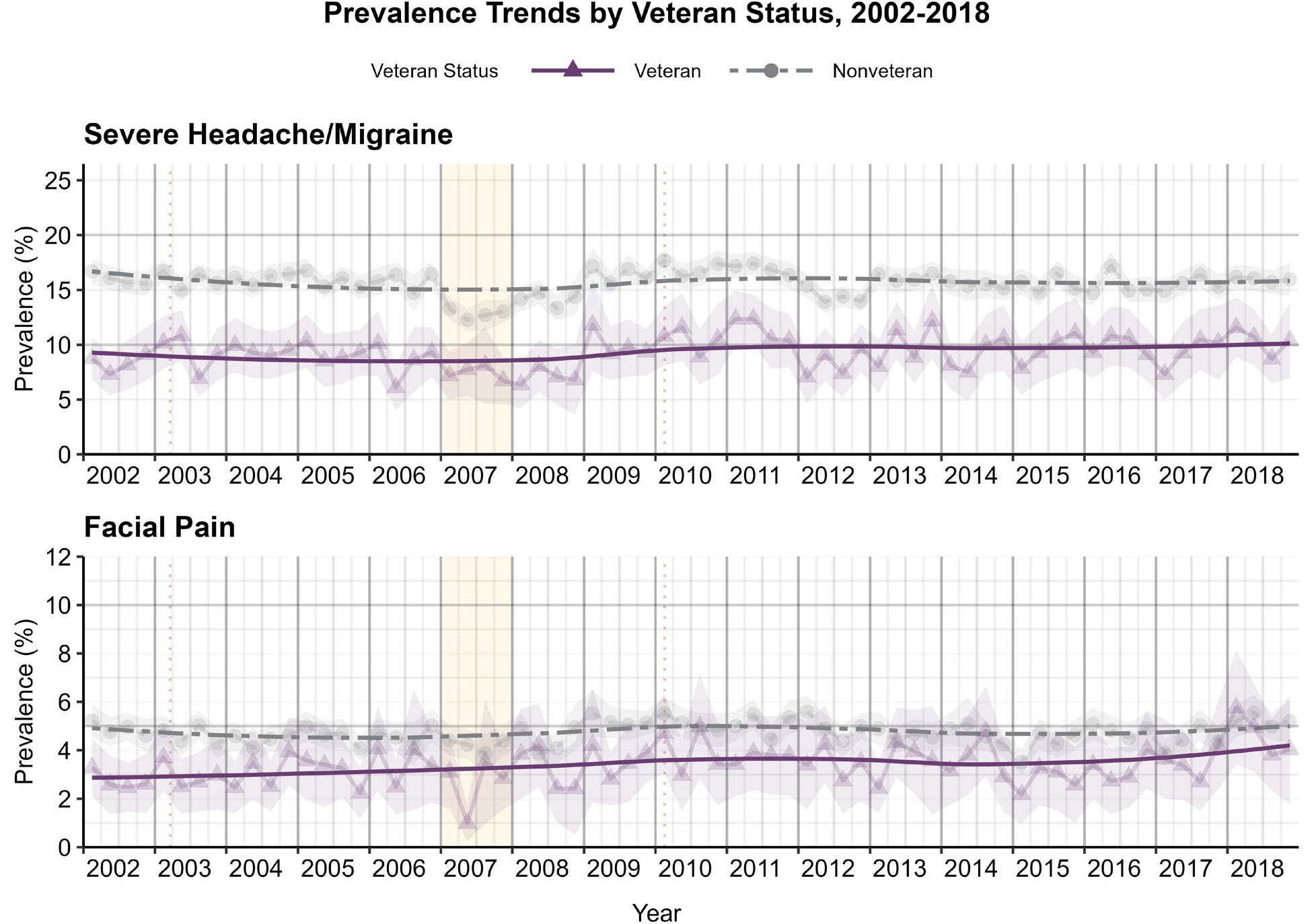
Prevalence Trends by Veteran Status, 2002-2018: Severe Headache or Migraine and Facial Pain. Annotations: Dotted lines represent initiation of Operation Iraqi Freedom in 2003 and Operation New Dawn in 2010. Yellow shading highlights in 2007, when common dips were seen across several pain variables.

### Facial Pain

Facial pain (**Figure 1**) had the lowest prevalence among both veterans and nonveterans. Like severe headache or migraine, crude facial pain prevalence was lower among veterans, ranging from 1.0% (95% CI: 0.3%, 1.7%; 2007, 2^nd^ quarter) to 5.8% (95% CI: 3.4%, 8.1%; 2018, 1^st^ quarter) among veterans and from 3.5% (95% CI: 3.0%, 4.1%; 2015, 1^st^ quarter) to 5.6% (95% CI: 4.9%, 6.3%; 2012, 1^st^ quarter) among nonveterans. Prevalence estimates and trend lines closely approximated when adjusting for demographic characteristics (**eFigure 1**).

### Neck Pain

Prevalence of neck pain (**Figure 2**) between group appear almost identical up until approximately 2009, where the prevalence trend for veterans appears to diverge and increase at a faster rate through the end of the study period. Crude neck pain prevalence ranged from 10.1% (95% CI: 6.6%, 13.6%; 2008, 4^th^ quarter) to 20.6% (95% CI: 16.5%, 24.7%; 2017, 4^th^ quarter) among veterans and from 12.8% (95% CI: 11.3%, 14.2%; 2017, 4^th^ quarter) to 17.6% (95% CI: 16.1%, 19.1%; 2018, 4^th^quarter) among nonveterans. Trend lines showed some separation when adjusting for demographic characteristics (**eFigure 2**), with slightly higher neck pain prevalence among veterans. Adjusted trend lines similarly diverged starting around 2009.

**Figure 2.**
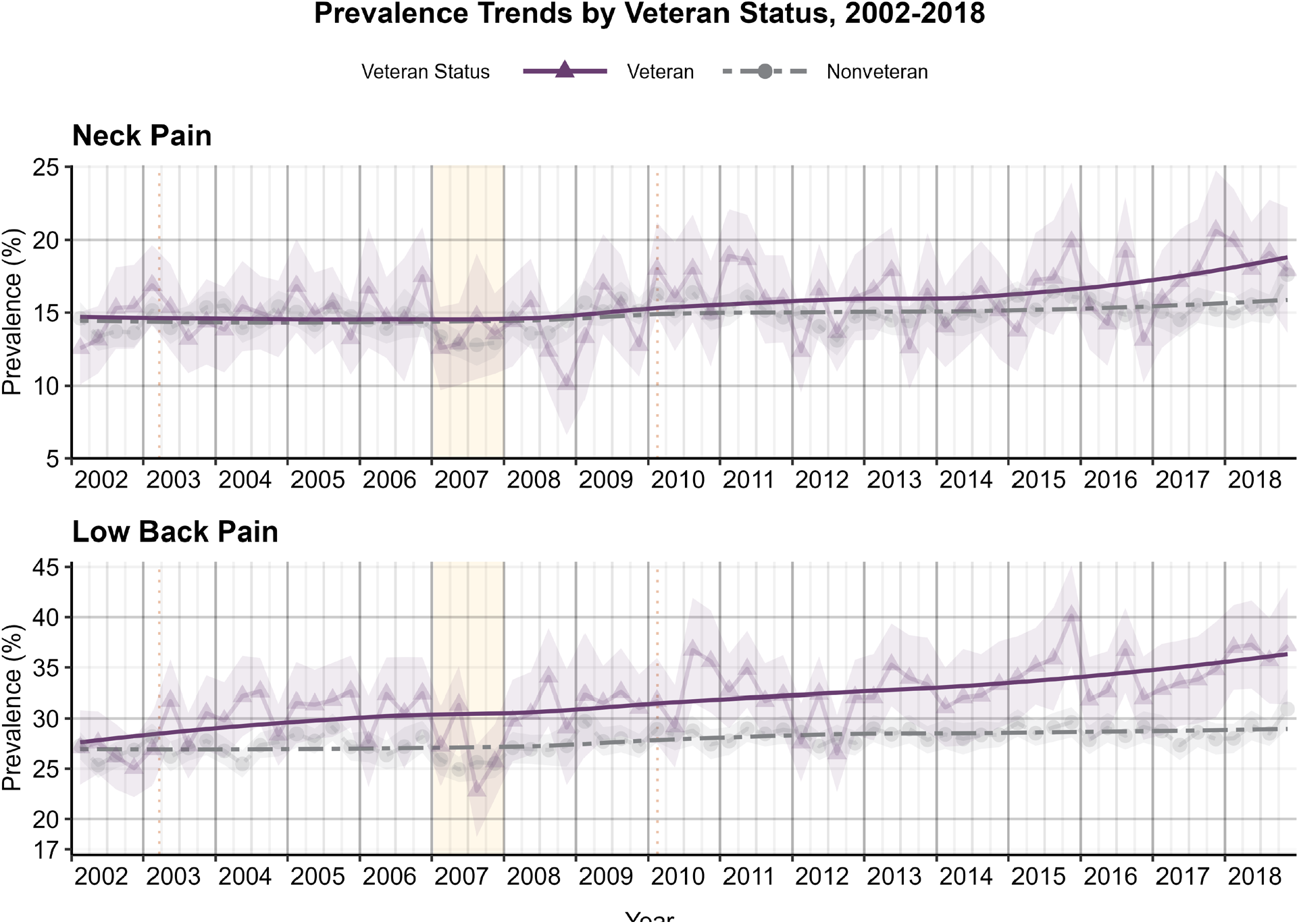
Prevalence Trends by Veteran Status, 2002-2018: Neck Pain and Low Back Pain. Annotations: Dotted lines represent initiation of Operation Iraqi Freedom in 2003 and Operation New Dawn in 2010. Yellow shading highlights in 2007, when common dips were seen across several pain variables.

### Low Back Pain

Prevalence of any LBP (**Figure 2**) was similar at the start of the study period; however, prevalence quickly began increasing, with trendlines diverging increasingly over time. Crude LBP prevalence ranged from 22.8% (95% CI: 18.2%, 27.4%; 2007, 3^rd^ quarter) to 40.2% (95% CI: 35.2%, 45.2%; 2015, 4^th^ quarter) among veterans and from 24.9% (95% CI: 23.7%, 26.6%; 2007, 2^nd^ quarter) to 30.9% (95% CI: 29.0%, 32.9%; 2018, 4^th^quarter) among nonveterans. When adjusting for demographic characteristics (**eFigure 2**), trend lines approximated but showed similar increasing trend divergence over time. We present results from LBP subgroup analyses in the **eResults, eFigure 3**, and **eFigure 4**.

### Joint Pain

Veterans had a notably higher crude joint pain prevalence (**Figure 3**) across the study period, with no overlap in prevalence estimates with nonveterans in any quarter. This ranged from 34.2% (95% CI: 29.9%, 38.5%; 2007, 1^st^ quarter) to 52.2% (95% CI: 46.7%, 57.8%; 2018, 4^th^ quarter) among veterans and from 25.6% (95% CI: 24.1%, 27.1%; 2007, 1^st^ quarter) to 34.2% (95% CI: 32.2%, 36.2%; 2018, 4^th^ quarter) among nonveterans. Joint pain prevalence estimates and trend lines closely approximated one another when adjusting for demographic characteristics (**eFigure 5**). These adjusted trends lines showed divergence near the end of the study period, with veterans starting to show slightly higher prevalence estimates.

**Figure 3.**
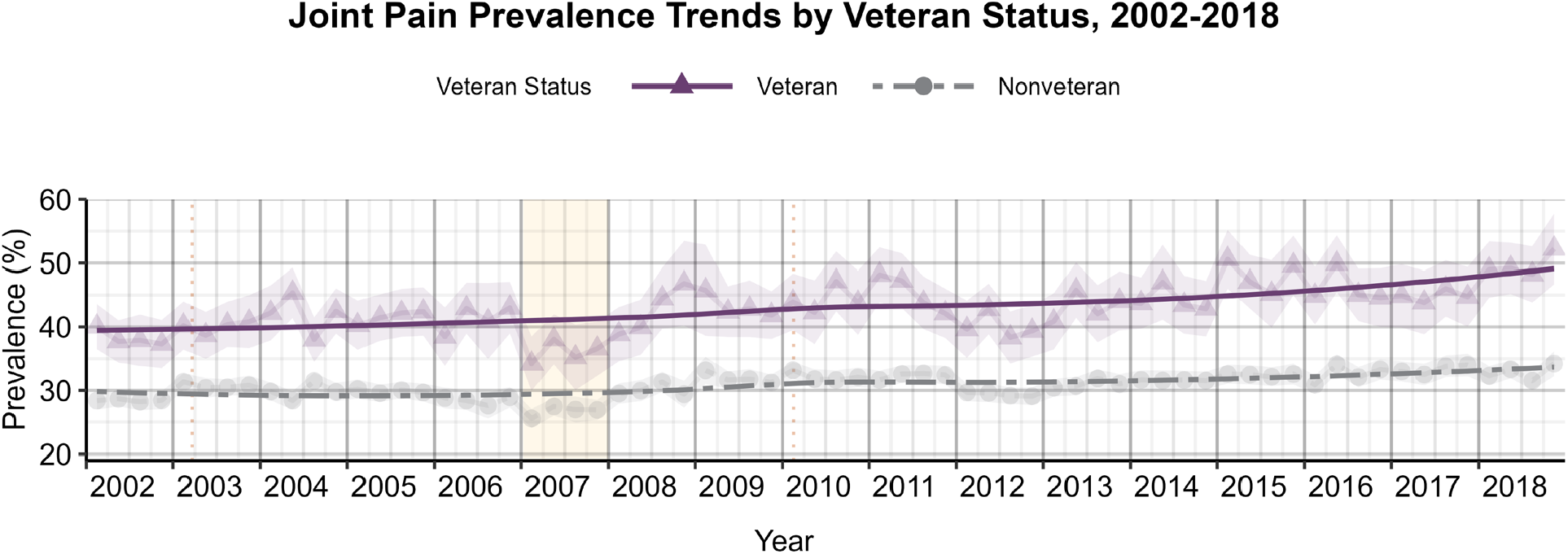
Joint Pain Prevalence Trends by Veteran Status, 2002-2018. Annotations: Dotted lines represent initiation of Operation Iraqi Freedom in 2003 and Operation New Dawn in 2010. Yellow shading highlights in 2007, when common dips were seen across several pain variables.

### Any Pain

Crude any pain prevalence (**Figure 4**) ranged from 49.2% (95% CI: 43.6%, 54.9%; 2007, 2^nd^ quarter) to 64.8% (95% CI: 59.3%, 70.2%; 2018, 4^th^ quarter) among veterans and from 45.9% (95% CI: 44.2%, 47.6%; 2007, 2^nd^ quarter) to 54.8% (95% CI: 52.5%, 57.1%; 2009, 1^st^ quarter) among nonveterans. Differences in any pain prevalence attenuated when adjusting for demographic characteristics (**eFigure 6**), but veterans continued to have higher prevalence than nonveterans across the study period.

**Figure 4.**
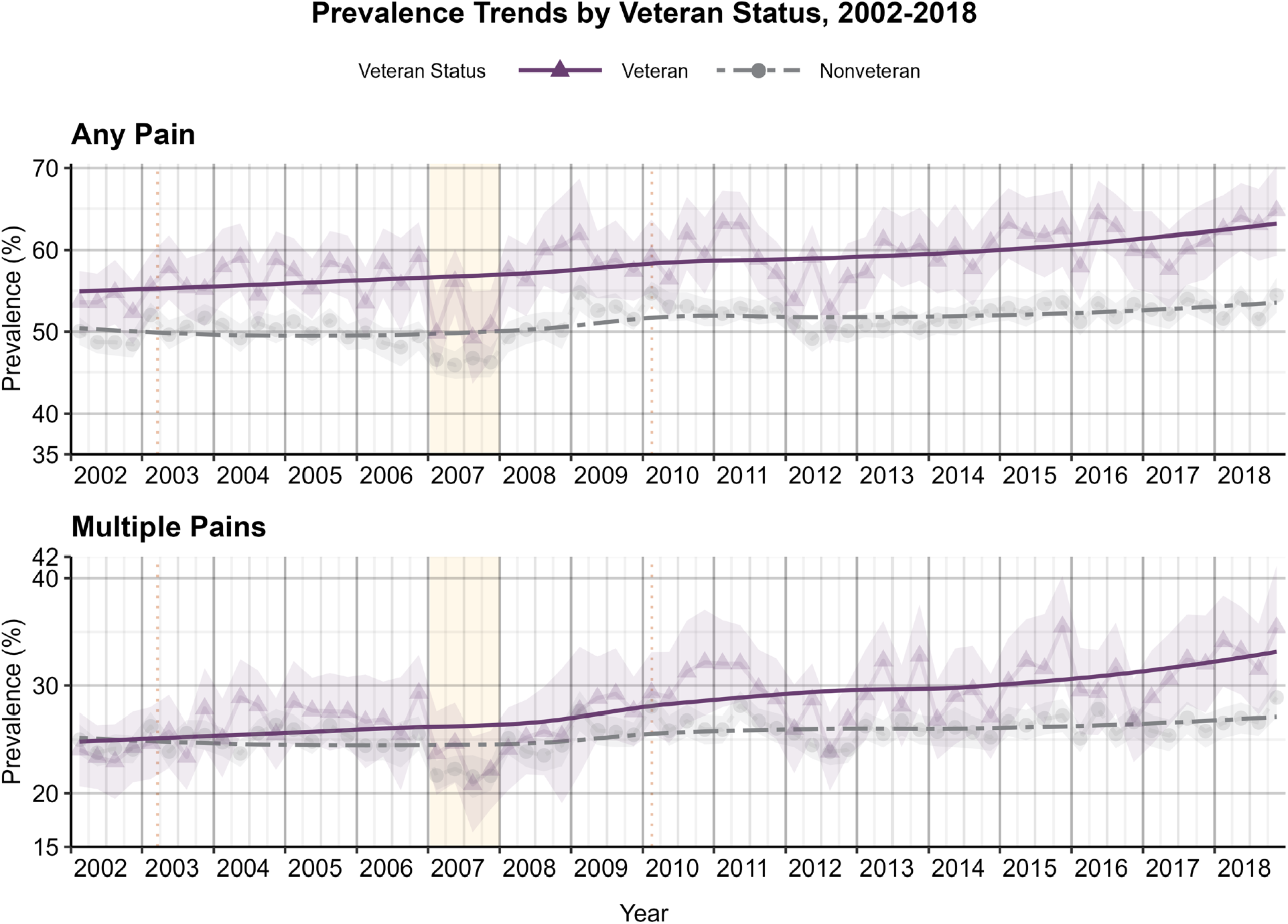
Prevalence Trends by Veteran Status, 2002-2018: Any Pain Complaint and Multiple Pain Complaints. Annotations: Dotted lines represent initiation of Operation Iraqi Freedom in 2003 and Operation New Dawn in 2010. Yellow shading highlights in 2007, when common dips were seen across several pain variables.

### Multiple Pains

Crude multiple pain prevalence (**Figure 4**) was similar between groups at the start of the study period; however, trends diverged around 2003 with higher prevalence of multiple pains among veterans across the second half of the study period. This ranged from 20.8% (95% CI: 16.4%, 25.3%; 2007, 3^rd^ quarter) to 35.5% (95% CI: 30.7%, 40.2%; 2018, 4^th^ quarter) among veterans and from 21.6% (95% CI: 19.6%, 23.6%; 2007, 3^rd^ quarter) to 28.9% (95% CI: 27.1%, 30.7%; 2018, 4^th^ quarter) among nonveterans. Trend lines showed more separation when adjusting for demographic characteristics (**eFigure 6**), with slightly more pronounced differences between veterans and nonveterans over time. Adjusted prevalence estimates showed similar increasing trend divergence starting around 2003.

We present the crude and adjusted mean differences in annual prevalence change between veterans and nonveterans in **Figure 5**.

**Figure 5.**
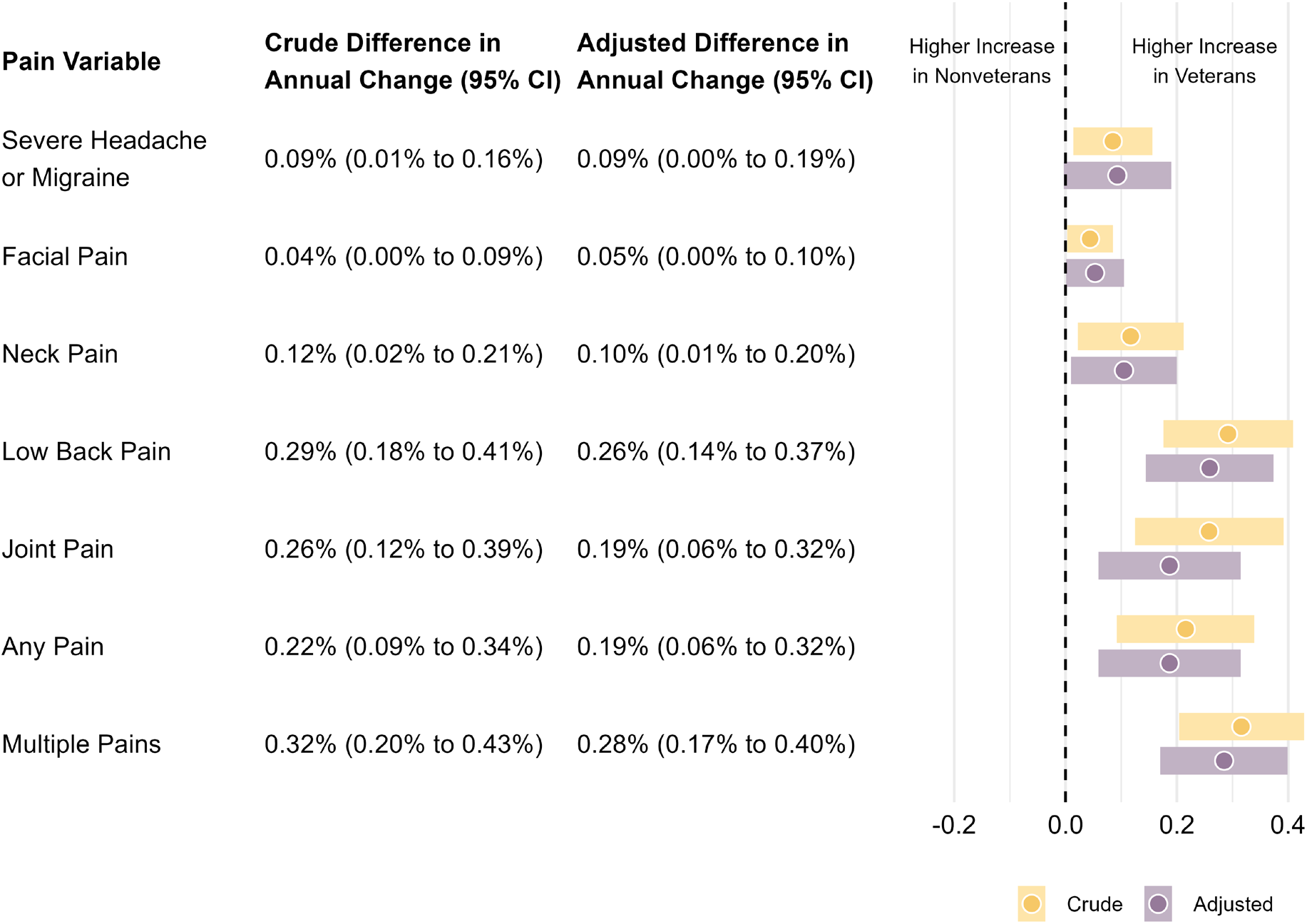
Mean Difference in Annual Change in Pain Prevalence Between Veterans and Nonveterans, 2002-2018. Annotations: Adjusted differences are adjusted for respondent-reported age, sex, race, and ethnicity.

## DISCUSSION

We investigated pain prevalence trends among U.S. military veterans compared to nonveterans from 2002 to 2018. The prevalence of most pain variables across the study period was either similar or higher among veterans compared to nonveterans. Further, all pain variables had larger prevalence increases across the 17-year study period among veterans, even after adjusting for demographic characteristics. These findings suggest that pain prevalence increased at a higher rate among veterans over this period.

Annual multiple pain prevalence grew 9.9% (41.7% relative increase) among veterans; an increase that was 3.3-times higher than nonveterans. Similarly, prevalence of having ≥1 of the 5 pain variables was higher among veterans, with annual prevalence increasing 10.0% (18.7% relative increase). This was 2.5-times higher than the increase among nonveterans.

Similarly, another study using 18 years of MEPS data reported an 8.1% absolute (25.0% relative) increase in the prevalence of any noncancerous painful health condition in the past year resulting in a medical event or disability episode.^11^ Our study builds onto this evidence by comparing veterans and nonveterans and capturing pain not limited by its connection to care-seeking or disability episodes.

Joint pain in veterans had the largest absolute increase in annual prevalence (11.4%); a 2.6-times higher total increase than nonveterans. A study of self-reported doctor-diagnosed osteoarthritis using nationally-representative data from the National Health and Nutrition Examination Survey (NHANES) years 2005-2018 also reported increasing age-adjusted prevalence trends.^27^ But, estimates were not presented by veteran status and reported prevalence was lower than our estimates of joint pain. The NHIS definition of joint pain in our study is not specific to doctor-diagnosed arthritis like in NHANES.

Neck and LBP trends in a study using all VHA data from 2004 to 2011 showed an increase from 1.9% to 2.5% (31.6% relative increase), 12.3% to 16.2% (31.7% relative increase), and 2.6% to 4.2% (61.5% relative increase) in veterans seeking care for neck, back, and spine pain in multiple spine segments, respectively.^14^ Prevalence of neck and LBP in our study were higher and had larger growth than trends from VHA data. These features are likely related to our study’s longer study period and more inclusive pain experiences that did not require a care-seeking encounter. Capturing pain regardless of VHA use in our study is a major strength, as only 30.4% of U.S. veterans used the VA for healthcare in 2019.^16^

Severe headache or migraine and facial pain had the lowest prevalence among veterans and – differently than other pain variables – were less prevalent than in nonveterans across the study period. Despite lower prevalence among veterans, our results showed larger increases in prevalence for severe headache or migraine (2.0% absolute increase; 24.2% relative increase) and facial pain (1.9% absolute increase; 69.4% relative increase) among veterans. In comparison, nonveterans had marginal changes over this same period. Interestingly, an increasing trend in migraine and non-migraine headaches has also been reported among active duty U.S. military personnel using military healthcare system data from 1998-2010, potentially coinciding with more military deployments from 2005-2010.^28^ When we adjusted for quarterly demographic characteristics, prevalence was slightly higher among veterans for both facial pain and severe headache or migraine. So, while fewer U.S. veterans have severe headache or migraine and facial pain, they appear to be slightly more burdened by these conditions than nonveterans when equalizing demographic characteristics. This reversal between crude and adjusted estimates is likely due to the greater proportion of males and higher mean age in the U.S. veteran population, given the strong female predominance (approximately 2:1) of severe headache or migraine and facial pain, and lower prevalence of these conditions after middle age.^28^

We observed notable dips in prevalence that occurred in 2007 among veterans for several pain variables. Nonveterans had a similar but less pronounced dip for some pain variables during the same year and demonstrated a similar 2007 dip in prevalence for severe headache and migraine not observed among veterans. The dips may be related to an NHIS sampling redesign implemented in 2006^21^ and budget shortfalls that reduced the targeted annual sample size from 2006-2008.^29^ The dips among veterans may be further related to a decrease of 458 thousand veterans in the total weighted population between 2006 and 2007. This reduction could be a result of voluntary (e.g., high reenlistment bonuses peaking in fiscal years 2006 and 2007) and involuntary military retention policies (i.e., Stop Loss) in place at that time.^30,31^ This timing also coincides with tour extension for many units (from 12 to 15 months) and escalation of troops overseas starting in 2007.^31-33^ However, the exact reasons for these dips are unclear. Adjusting prevalence for quarterly demographic characteristics attenuated veteran and nonveteran trendline differences to varying degrees for severe headache or migraine, facial pain, joint pain, and LBP. On the other hand, demographic adjustment for neck pain and multiple pain prevalence resulted in a slight separation of trends.

There are no known direct comparisons of healthcare utilization trends for pain between veterans (regardless of their VHA use) and nonveterans. Our study addresses this gap, comparing trends in several pain variables in a representative sample of these subgroups of the U.S. adult population. Prior studies indicate that pain is one of the most expensive health conditions in the U.S.^34,35^ Thus, we expect that if the disparate upward trends in pain prevalence were to continue, they would likely come with similar disparate trends in associated healthcare needs. This concern is compounded by evidence that veterans experience higher pain severity than nonveterans^10^, which is associated with higher healthcare utilization.^36^

### Limitations

Our study provides estimates representative of the overall non-institutionalized U.S. veteran population, receiving care within and outside of the VHA. While this is a strength, VHA enrollment status of veteran respondents is unknown for our study period because the NHIS did not collect this variable before 2019. Future studies using data from the NHIS 2019 redesign and beyond will be able to investigate these veteran subgroups further. Likewise, the NHIS does not collect additional military service characteristics from veterans that may be of interest (e.g., combat exposure, number and duration of deployments). The NHIS excludes older individuals who are living in long-term care institutions, adults of any age living in correctional facilities, and U.S. nationals living abroad; limiting generalizability to those veterans. Pain questions used in our study asked respondents about pain during the past 3 months (or 30 days for joint pain). The wording limits determination of chronicity, intensity, or interference or persistence. Further, the NHIS is an annually repeated cross-sectional survey; although the target population of interest is the same across all years, the NHIS does not follow respondents longitudinally, preventing individual-level inferences from year-to-year.

## CONCLUSIONS

This study is the first to report trends in pain prevalence among veterans outside of the VHA that are directly comparable to nonveterans in terms of representativeness and pain variable ascertainment. Both absolute and relative total increases for all pains were larger among veterans than nonveterans over the 17-year study period. Veterans had higher rates of increase over time and had similar or higher prevalence of all pain variables compared to nonveterans with similar demographic characteristics. Continued accelerated increase in pain among veterans would likely impact healthcare utilization (within and outside of the VHA) and underscores the need for improved pain prevention and care programs for these individuals with disproportionate pain burden.

## Data Availability

Data used in this study are freely available for download from IPUMS Health Surveys (https://nhis.ipums.org/). SAS Code used for analysis is available via GitHub (https://github.com/KennethATaylor/Veteran-Pain-Trends).

https://github.com/KennethATaylor/Veteran-Pain-Trends

## Author Contributions

Dr. Taylor and Dr. Kosinski had full access to all of the data in the study and take responsibility for the integrity of the data and the accuracy of the data analysis.

*Concept and design:* Taylor, Goode.

*Acquisition, analysis, or interpretation of data:* Taylor, Kapos, Kosinski, Sharpe, Rhon, Goode.

*Drafting of the manuscript:* Taylor, Kapos, Sharpe, Rhon, Goode.

*Critical revision of the manuscript for important intellectual content:* Taylor, Kapos, Kosinski, Sharpe, Rhon, Goode.

*Statistical analysis:* Taylor, Kosinski.

*Obtained funding:* Goode.

*Administrative, technical, or material support:* Taylor.

*Supervision:* Taylor, Goode.

## Conflict of Interest Disclosures

Dr. Taylor and Dr. Goode report receiving grant funding from National Institute of Arthritis and Musculoskeletal and Skin Diseases (NIAMS), during the conduct of the study. No other disclosures were reported.

### Disclaimer

The findings and conclusions in this report are those of the authors and do not necessarily represent the official position of NIAMS.

## Additional Contributions

